# Health care use up to 6 months after COVID-19 in 700.000 children and adolescents: a pre-post study

**DOI:** 10.1101/2021.06.02.21258211

**Authors:** Karin Magnusson, Katrine Damgaard Skyrud, Pål Suren, Margrethe Greve-Isdahl, Ketil Størdal, Doris Tove Kristoffersen, Kjetil Telle

**Author notes:** **Corresponding author, contact information:** Karin Magnusson, Cluster for Health Services Research, Norwegian Institute of Public Health, Postboks 222, Skøyen, N-0213 Oslo. Visiting address: Sandakerveien 24c, Building D, 0473 Oslo.

## Abstract

**Objectives:** To explore whether, and for how long COVID-19 among children gives an increase in use of health care services, when compared to children with no COVID-19.

**Methods:** Studying all Norwegian residents aged 1-5, 6-15 and 16-19 years from August 1^st^ 2020 to February 1^st^ 2021 (N= 768 560), we contrasted rates of monthly all-cause primary and specialist health care use before and after testing for SARS-CoV-2 (% relative change), for children testing positive (non-hospitalized in the acute phase) (N=10 306) vs children with no COVID-19 (N=758 254).

**Results:** We found a substantial elevation in short-term primary care use for children testing positive for SARS-CoV-2 during the first month following positive test when compared to children testing negative (relative elevation 1-5 years: 325%, 95%CI=296-354; 6-15 years: 434%, 95%CI=415-453; 16-19 years: 360%, 95%CI=342-379). There was still elevated primary care use at 2 months (1-5 years: 21%, 95%CI= 4-38; 6-15 years: 13%, 95%CI=2-25) and at 3 months (1-5 years: 26%, 95%CI=7-45, 6-15 years: 15%, 95%CI=3-26) for young children, but not at 2 or 3 months for the older children (16-19 years: 10%, 95%CI=-1-22 and 6%, 95%CI=-5-18, respectively). The 1-5-year-olds also had a long-term (up to 6 months) increase of primary care (14%, 95%CI=1-26) that was not observed for older age groups, when compared to same-aged children testing negative. We observed no elevated use of specialist care.

**Conclusion:** Children in pre-school age used health services for a longer time (3-6 months) after COVID-19 than children in primary and secondary school age (1-3 months).

## Introduction

The content, duration and impact of the post-covid syndrome has been described for adults in several studies. We recently showed that adults with severe COVID-19 may experience complaints for up to 3-6 months after initial infection, mainly due to respiratory and circulatory conditions^1^. These findings line up with a range of other reports showing increased risk of complications after serious disease among adults, implying that severe COVID-19 has a considerable impact on long-term health care utilization^2-5^. Less is known about potential long-term sequela after *mild* COVID-19^5^.

Except for the occurrence of the rare multisystem inflammatory syndrome in children (MIS-C) after initial COVID-19^6^ and a good prognosis in terms of death^7^, far less is known about the impact of COVID-19 on post-covid health and health care use among children in general. Existing studies are case-reports and analyses of small populations including from 5 to 33 children, mostly severely affected by initial COVID-19, with similar long-term complaints observed in children as in adults^8, 9^. In one of the largest studies of children with COVID-19 to date, more than half of 129 children with mean age 11 years reported at least one persisting symptom 120 days after COVID-19^10^. This contrasts another of the largest studies, reporting that only 4% of 171 children with median age 3 years had persistent symptoms at 3-8 weeks following the initial infection^11^. Neither of these studies are useful in the development and implementation of policy to face the pandemic.

Large-scale, population-based and prospective studies are definitely needed to determine the magnitude, duration and impact of long-lasting complaints following COVID-19 among children. Also, little is known whether health care use among children with COVID-19 is increased after the initial disease at different ages, and for how long the increase would persist for young vs old children. As an example, a long-lasting increase in specialist care visits following COVID-19 would imply that the post-covid complaints are severe for a certain age group, whereas a short increase restricted to primary care, would imply milder complaints. Such knowledge may be used to upscale or downscale the health services. We aimed to explore the short term (0-3 months) and long term (4-6 months) impacts of potentially long-lasting complaints following COVID-19 which resulted in health care utilization among children and adolescents aged 1-5, 6-15 and 16-19 years.

## Methods

### Design & data sources

To estimate the long-term impacts of COVID-19 on health care use among children we utilized population-wide longitudinal registry data from Norway. The BeredtC19 register is an emergency preparedness register aiming to provide rapid knowledge about the pandemic, including impacts of measures to limit the spread of the virus on health and utilization of health care services^12^. BeredtC19 compiles daily updated individual-level data from several registers, including the Norwegian Surveillance System for Communicable Diseases (MSIS) (all testing for COVID-19), the Norwegian Patient Register (NPR) (all electronic patient records from all hospitals in Norway), and the Norway Control and Payment of Health Reimbursement (KUHR) Database (all consultations with all general practitioners and emergency primary health care) as well as the National Population Register (age, sex, country of birth, date of death). Thus, BeredtC19 includes all polymerase chain reaction (PCR) tests for SARS-CoV-2 in Norway with date of testing and test result, reported from all laboratories in Norway to MSIS and all electronic patient records from primary care as well as hospital-based outpatient and inpatient specialist care. The establishment of an emergency preparedness register forms part of the legally mandated responsibilities of The Norwegian Institute of Public Health (NIPH) during epidemics. Institutional board review was conducted, and The Ethics Committee of South-East Norway confirmed (June 4th 2020, #153204) that external ethical board review was not required.

### Study population

Our population included all residents of Norway aged between 1 and 19 years on January 1^st^ 2020, and who had been tested for the SARS-CoV-2 by a PCR-test, with a positive or negative test result. We also included all untested children, and randomly assigned them a hypothetical test date. To account for temporal changes in testing intensity, the hypothetical test date was assigned so that the fraction of untested children with a test date in each calendar week was the same as the fraction of tested children with an actual test date in the same calendar week. Since testing intensity differed by age groups over time, this assignment was done separately for age group 1-5 years (pre-school age), 6-15 years (primary and lower secondary school age) and 16-19 years (upper secondary school age). In the beginning of the pandemic (March-July 2020) children were less frequently tested (i.e. not included in test criteria), and we therefore restricted to the time period from August 1^st^ 2020 to February 1^st^ 2021, when PCR tests were widely available and the 2^nd^ wave of infection was at its beginning in Norway.

Also, because parents may be more likely to test children with pre-existing conditions than healthy children, and because many tests were performed as a routine prior to hospital visits, patients receiving hospital-based outpatient or inpatient specialist care during the test week, and during the first or second week following the test week, were excluded. For consistency, we used similar exclusion criteria for the group of untested children. Note that this requirement implies that we do not include the few children who were hospitalized (inpatient and outpatient) because of serious initial COVID-19 in our study. With outcome data from May 1^st^ 2020 through May 1^st^ 2021, we could follow children in the different age groups from three months before to at least three months after the test date.

### COVID-19

We studied all children who were tested and not tested for SARS-CoV-2 in Norway, divided into three mutually exclusive groups:

1. COVID-19, comprising all children with one or more positive PCR tests (using the first available test date with a positive result) and not admitted to hospital with COVID-19.
2. No COVID-19, comprising all children with one or more negative PCR tests (comparison group 1), choosing a random test date for children with several tests with negative results.
3. Untested, comprising all children with no PCR test, who were randomly assigned a hypothetical test date (comparison group 2).

### Outcomes

We studied all-cause health care use in 1) primary and 2) specialist care from three months before to six months or more after the test week. The categorical outcome variable for primary care was set to one if the child had visited *primary care* (i.e. general practitioners or emergency wards) at least once during a week (otherwise zero), and the categorical outcome variable for specialist care to one if the child had received hospital-based *outpatient- or inpatient specialist care* at least once during a week (otherwise zero). Observations for children who had a short follow-up time (i.e. who were tested December 2020 –February 2021) were censored from the week that we could no longer observe health care use.

### Statistical analyses

We first explored descriptive data by age group, such as testing patterns (percentage having at least one PCR-test and percentage with a positive test among the tested), sex, immigrant background and comorbidities. Second, we studied the percent using health care services at least once per week from 3 months prior to test week, to 6 months after test week for the children with COVID-19, without COVID-19 and the untested children, by age group. Thus, we calculated the percent using health services per calendar week, and presented averages over the 3 months before test week, and over post-test periods 1-4 weeks, 5-8 weeks, 9-12 weeks and 13-24 weeks. We also plotted the percentages by periods of four weeks following the test week (these were adjusted for potential confounders as described beneath).

Third, to estimate how much larger or smaller the use of health care services was for children with COVID-19 compared with 1) children with no COVID-19 or 2) untested children, we used a generalized difference-in-differences (DiD) approach. DiD analysis evaluate the effect of an event by comparing the change in the outcome for the affected group before and after the event, to the change over the same time span in a group not affected by the event^13-15^. In this study, we compared the rate of health care use in the months before and after the PCR test for the children who tested positive (difference 1), to the difference in the rate of health care use in the months before and after the PCR test for 1) the children with no COVID-19 or 2) the untested children (difference 2). The DiD estimate is the difference between these two differences, estimated using linear probability models with robust standard errors and presented as a difference in percentage points. Statistically, one uses an interaction term (between pre-post PCR-tests and COVID-19 group category) to derive the DiD estimate. By including calendar month fixed effects, this approach accounts for background trends like seasonal variations in health care use^13^. The DiD estimate can be interpreted as the change in health care use that is related to COVID-19, beyond any background calendar month trends. If there is no relationship between COVID-19 and subsequent health care use, the DiD estimate would be zero.

We generalized this traditional DiD method by extending from one to four post-test periods: 1-4 weeks, 5-8 weeks, 9-12 weeks and 13-24 weeks, comparing to one pre-period (the 3 months before test week) and also including a separate parameter for the test week. The generalization was implemented by including categorical variables for each of these extra periods and accompanying interaction terms. In addition to the presentation of results as absolute differences in percentage points, we also presented relative differences (i.e. in percent) by dividing the absolute estimate (and corresponding lower and upper confidence interval bounds) for each of the post periods by the health care use rate of the comparison group in the pre period (and multiply by 100).

DiD models are used in two data situations, one where *different* individuals are studied before and after the event, and another, which is our case, where the *same* individuals are followed from before to after the event^14^. While adjusting for individual characteristics that are constant over time can be important in the first type of data situation to account for changes in composition, it is less likely to affect our DiD estimates where the same individuals are followed over time. However, composition might also matter in our situation due to the censoring, and adjusting may improve precision^14^. We thus adjusted for the following individual characteristics: Sex (boys/girls), comorbidities (categories 0, 1, 2 or 3 or more comorbidities) based on risk conditions for COVID-19 defined by an expert panel^16^, birth country (Norway/abroad) and calendar month (12 categories).

Models were run separately for each of the age groups 1-5 years, 6-15 years and 16-19 years, as well as for the two outcomes all-cause health care use in primary care, and all-cause health care use in specialist care. For the outcomes that showed elevated health care use following positive test for SARS-CoV-2, we also repeated the analyses using cause-specific outcomes as described in the Supplementary (S-)Table 1 (causes: digestive, circulatory, respiratory, endocrine/metabolic/nutritional, genitourinary, eye/ear, musculoskeletal, mental, skin, blood and general/unspecified conditions). All analyses were run in STATA MP v.16.

**Table 1.** Descriptive characteristics.

|  | Age 1-5 years |  |  | Age 6-15 years |  |  | Age 16-19 years |  |  |
| --- | --- | --- | --- | --- | --- | --- | --- | --- | --- |
|  | COVID-19 | No COVID-19 | Unt. Contr. | COVID-19 | No COVID-19 | Unt. Contr. | COVID-19 | No COVID-19 | Unt. Contr. |
| Population, n | 1362 | 45113 | 115538 | 5585 | 161612 | 283966 | 3690 | 90977 | 90718 |
| Hospitalized^<br>(excluded), n (%) | 25 (1.8) | 3802 (8.4) | 2680 (2.3) | 158 (2.8) | 10402 (6.4) | 10122 (3.6) | 148 (4.0) | 6566 (7.2) | 3699 (4.1) |
| Study sample, non-hospitalized^<br>(included), N | 1337 | 41311 | 112858 | 5427 | 151210 | 273844 | 3542 | 84411 | 87019 |
| Age, mean (SD) | 3.2(1.4) | 3.0(1.4) | 3.1(1.4) | 11.1(2.8) | 11.2(2.8) | 10.2(2.8) | 17.6(1.1) | 17.5(1.1) | 17.5(1.1) |
| Girls, % | 47.1 | 46.8 | 49.0 | 48.9 | 48.5 | 48.9 | 48.3 | 50.5 | 44.5 |
| Born abroad*, % | 6.3 | 3.1 | 7.0 | 15.9 | 7.6 | 14.5 | 21.6 | 10.43 | 18.1 |
| ≥2 comorbidities, % | 0.15 | 0.12 | 0.05 | 0.24 | 0.21 | 0.09 | 0.11 | 0.21 | 0.13 |

^Inpatient or outpatient during test week or 1-2 weeks after (hypothetical) test week. \*Based on child's own country of birth.

## Results

Of the about 1.3 million (N=1 289 991) children aged 1-19 years living in Norway, we studied all children aged 1-19 years (N= 768 560) who had been tested (N= 294 839) or who were not tested (N= 473 721) for SARS-CoV-2 in Norway from August 1^st^ 2020 to February 1^st^ 2021. In total, 10 306 children tested positive (1.3% of the study population). The 521 431 children who were excluded were mainly children who had their (hypothetical) test date prior to August 1^st^ 2020 or after February 1^st^ 2021 (N=483 829), or children who were hospitalized (inpatient or outpatient) during the test week or the 1^st^ or 2^nd^ week following the (hypothetical) test week (N=37602) (Table 1).

Figure 1 shows that, on average, over all months, children in the youngest age group (1-5 years) were tested somewhat less frequently than children in the oldest age group (16-19 years). However, the percentage testing positive was more or less similar for the different age groups and increasing throughout the study period (Figure 1). Table 1 shows that the children testing positive for SARS-CoV-2 and the two comparison groups were similar in terms of age and sex. However, children testing negative were more often Norwegian-born compared to untested children (with their randomly assigned test date, see above) and compared to children testing positive, and a larger proportion among children being tested had 2 or more comorbidities (Table 1).

**Figure 1.**
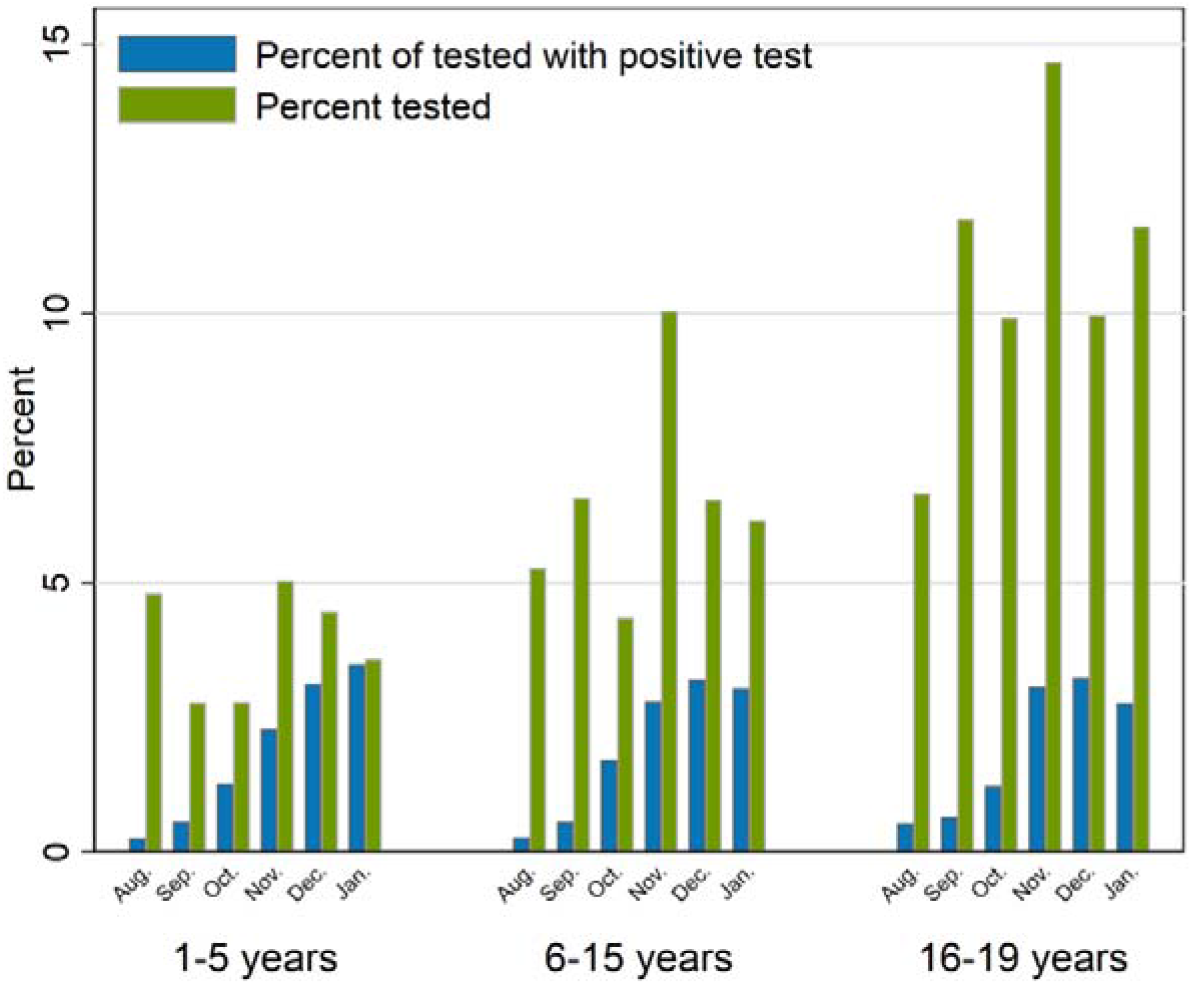
Percentage of the sample being tested for SARS-CoV-2 in PCR test by month (August to December 2020, and January 2021), and percentage of the tested who tested positive for SARS-CoV-2 per month, by age group.

### Group-wise change in rates of health care use following test week

In total 3.3% of children aged 1-19 years with positive SARS-CoV-2 test used *primary care* in the three months before being tested, which increased to 41% in the test week, before declining steadily by weeks 1-4 weeks (16%), 5-8 weeks (3.9%), 9-12 weeks (3.9%) and 13-24 weeks (2.9%) after test week (Table 2). Thus, by weeks 9-12 and 13-24, the proportion utilizing primary care following positive test had declined to more or less similar levels as were observed prior to the positive test. For children testing negative, the increase in the test week was less steep and more rapidly went back to pre-test levels (Table 2). For the untested children, the utilization of primary care was very similar from 3 months before to 6 months after the (hypothetical) test week (Table 2).

**Table 2.**
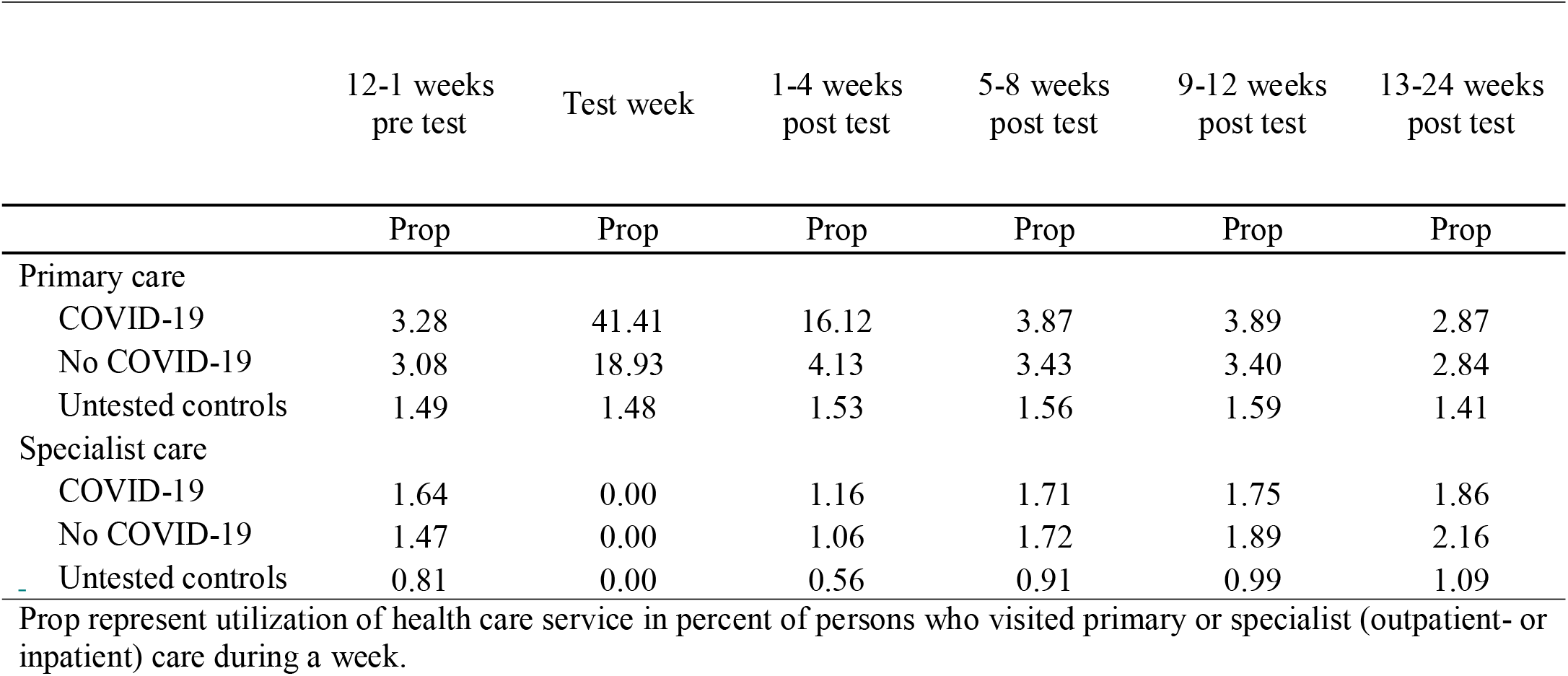
Utilization of health care services in given time periods before and after PCR test for SARS-CoV-2, in percent of children who visited primary or specialist (outpatient- or inpatient) care during a week, separately for those with COVID-19, no COVID-19 and untested controls.

No increased rates of *specialist care* were observed among the children testing positive for SARS-CoV-2, nor among the children testing negative or who were untested (Table 2). Since we only studied children with mild disease courses of initial COVID-19 (i.e. children who were not hospitalized (inpatient or outpatient) in the test week or in the 1 or 2 weeks following the test week), the zero specialist consultations observed in the test week (Table 2) occur by deliberate construction of the study population.

Similar patterns, i.e. with a steep rise in primary, but not specialist care use following a positive test for SARS-CoV-2, were confirmed in age-specific plots (1-5, 6-15 and 16-19 years) that were adjusted for sex, birth country, comorbidities and calendar month (Figure 2).

**Figure 2.**
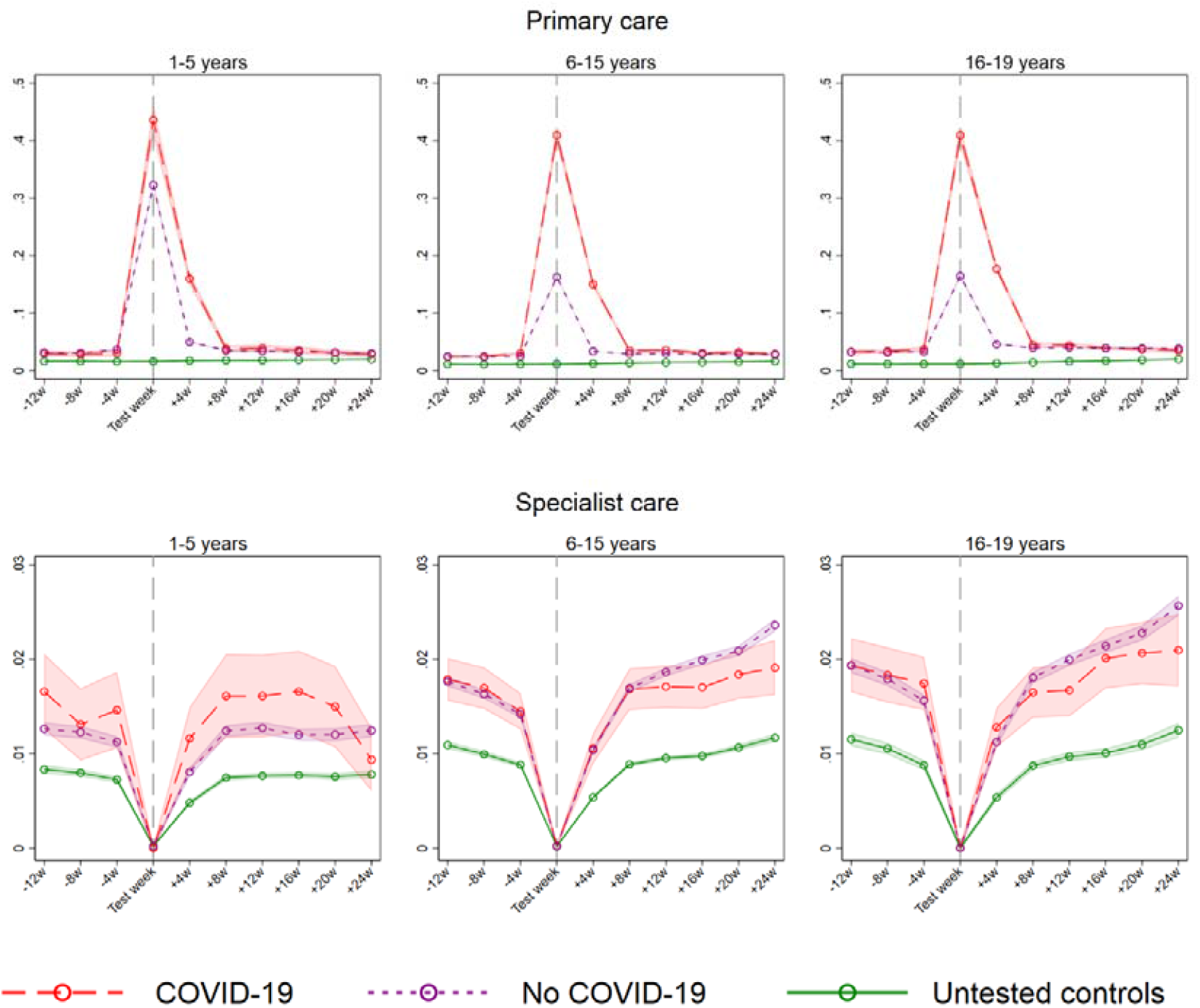
Estimated percent of children using (95% CI) primary and specialist (inpatient and outpatient) care in a week, from 3 months before to 6 months after week of PCR test for SARS-CoV-2, for COVID-19, no COVID-19 and untested, by age groups. Estimates adjusted for age, sex, comorbidities, birth country and calendar month. The dip for specialist care around the test week is a mechanical result of the exclusion of children who were hospitalized with COVID-19 in the test week and the two following weeks.

### Short term (0-3 months) effects on health care when compared to children with no COVID-19

When applying the DiD models and statistically comparing the within-group changes over time with each other, we observed a substantial short-term increase in primary care use for children testing positive for SARS-CoV-2 during the first 1-4 weeks following positive test when compared to children testing negative (age 1-5: 325%, age 6-15: 434% and age 16-19: 360% relative increase) (Table 3). At 5-8 weeks post-test, there was still an increase in primary care use of ∼13-21% for children in age groups 1-5 and 6-15 who tested positive, when compared to children testing negative (Table 3). However, when comparing the oldest children testing positive, to the oldest children testing negative at 5-8 weeks, the absolute DiD estimate was not significantly different from zero (B=0.38, 95% CI= -0.04-0.79) implying no group difference in change in primary care use over time for the oldest children (Table 3).

**Table 3.** Impacts of COVID-19 on health care utilization in children, using children testing negative as comparison group. Differences-in-differences estimates (in percentage points) for the change in the rate of persons utilizing the health care service in question per week, after the week of PCR test for SARS-CoV-2.

|  | Primary care |  | Specialist care |  |
| --- | --- | --- | --- | --- |
|  | B (95% CI) | % rel. diff.<br>(95 % CI) | B (95% CI) | % rel. diff. (95 % CI) |
| Age 1-5 years |  |  |  |  |
| 1-4w | 11.46 (10.43,12.49) | 325 (296,354) | 0.11 (-0.26,0.48) | 3 (-7,14) |
| 5-8w | 0.75 (0.16,1.4) | 21 (4,38) | 0.12 (-0.32,0.56) | 3 (-9, 16) |
| 9-12w | 0.91 (0.26,1.57) | 26 (7,45) | 0.09 (-0.35,0.53) | 3 (-10,15) |
| 13-24w | 0.48 (0.03,0.3) | 14 (1,26) | -0.07 (-0.42,0.28) | -2 (-12,8) |
| Age 6-15 years |  |  |  |  |
| 1-4w | 11.57 (11.06,12.08) | 434 (415,453) | 0.02 (-0.17,0.21) | 1 (-6, 8) |
| 5-8w | 0.36 (0.05,0.66) | 13 (2,25) | -0.008 (-0.23,0.22) | 0 (-9,8) |
| 9-12w | 0.39 (0.09,0.70) | 15 (3,26) | -0.15 (-0.39,0.09) | -6 (-15,3) |
| 13-24w | 0.04 (-0.18,0.25) | 1 (-7,9) | -0.32 (-0.54,-0.09) | -12 (-20,-4) |
| Age 16-19 years |  |  |  |  |
| 1-4w | 13.01 (12.33,13.69) | 360 (342,379) | 0.11 (-0.14,0.36) | 3 (-410) |
| 5-8w | 0.38 (-0.04,0.79) | 10 (-1,22) | -0.21 (-0.48,0.061) | -6 (-13, 2) |
| 9-12w | 0.23 (-0.19,0.64) | 6 (-5,18) | -0.38 (-0.68,-0.08) | -10 (-19,-2) |
| 13-24w | -0.30 (-0.58,-0.01) | -8 (-16,-0) | -0.32 (-0.63,-0.02) | -9 (-17,0) |

Beta (B) estimates (95% confidence intervals - CI) represent the difference in the weekly rate (in percentage points) of health care utilization during 1-4 weeks, 5-8 weeks, 9-12 weeks and 13-24 weeks compared with during 12-1 weeks before, for those testing positive for COVID-19 vs children testing negative for COVID-19. Estimates compare the post-test period (as indicated) to the pre-test period (12-1 weeks before) for those testing positive for COVID-19, to the same pre- and post-periods for those testing negative for COVID-19.
Categorical variables for utilization in test week, age, sex, calendar month, birth country and comorbidities were included in all models. The % relative diff. transforms the difference-in-difference estimate, which are in percentage points, to the relative change compared with the utilization rate during 12-1 weeks before the PCR test for the group with no COVID-19 (with lower and upper bound of CI also calculated by dividing by the utilization rate during 12-1 weeks before the PCR test for the group with no COVID-19). PCR: polymerase chain reaction.

The tendency towards younger children having a lengthier increase in health care use following COVID-19, was again observed in group-wise comparisons of health care use at 9-12 weeks after testing. Here, the children aged 1-5 and 6-15 years who tested positive, had a 26% and 15% relative increase in primary care use when compared to same-aged children testing negative, respectively (Table 3). Children aged 16-19 with positive test had no elevated primary care use at 9-12 weeks when compared to same-aged children testing negative (Table 3). For all age groups, complaints from the respiratory system, as well as general and unspecified conditions, were the main causes for the elevated primary care use (S-Figure 1).

We observed no short-term increase in *specialist care* use for any of the age groups following a positive test for SARS-CoV-2, neither at 1-4, 5-8 or 9-12 weeks when compared to children testing negative (Table 3).

### Short term (0-3 months) effects on health care when compared to untested children

A similar pattern was observed when repeating the analyses using untested controls who were assigned a hypothetical test date, although the short-term differences in group-wise change were generally larger than when using children testing negative as a control group (Table 3, Table 4). Again, we observed a large immediate increased primary health care use during the first month following positive test, for all age groups (695% relative increase for children aged 1-5 years, 911-919% relative increase for children aged 6-19 years) (Table 4). In the 5-8 weeks as well as 9-12 weeks following, the increase was still considerable, i.e. children in *all* age groups who tested positive, had an increased primary health care relative to the untested children in the same age group (relative increase 33-49%) (Table 4). Complaints from the respiratory system, and general and unspecified conditions were the main causes for the increased primary care use for all post-covid periods, for all age groups (S-Figure 1).

**Table 4.** Impacts of COVID-19 on health care utilization in children, using untested controls as comparison group. Differences-in-differences estimates (in percentage points) for the change in the rate of persons utilizing the health care service in question per week, after the week of PCR test for SARS-CoV-2.

|  | Primary care |  | Specialist care |  |
| --- | --- | --- | --- | --- |
|  | B (95% CI) | % rel. diff. (95 % CI) | B (95% CI) | % rel. diff. (95% CI) |
| Age 1-5 years |  |  |  |  |
| 1-4w | 12.85 (11.82,13.87) | 695 (640,750) | -0.022 (-0.38,0.34) | -3 (-54,48) |
| 5-8w | 0.61 (0.028,1.2) | 33 (2,65) | 0.16 (-0.27,0.6) | 23 (-39,84) |
| 9-12w | 0.68 (0.03,1.3) | 37 (1,72) | 0.14 (-0.30,0.58) | 20 (-42,81) |
| 13-24w | -0.11 (-0.55,0.33) | -6 (-30,18) | -0.12 (-0.46,0.23) | -16 (-65,33) |
| Age 6-15 years |  |  |  |  |
| 1-4w | 12.20 (11.70,12.71) | 911 (873,948) | -0.16 (-0.35,0.03) | -19 (-42,4) |
| 5-8w | 0.59 (0.29,0.89) | 44 (22,67) | 0.11 (-0.11,0.33) | 14 (-13,40) |
| 9-12w | 0.66 (0.36,0.96) | 49 (27,71) | 0.07 (-0.17,0.31) | 8 (-20,37) |
| 13-24w | -0.03 (-0.24,0.18) | -2 (-18,14) | 0.05 (-0.17,0.27) | 6 (-20,32) |
| Age 16-19 years |  |  |  |  |
| 1-4w | 14.00 (13.32,14.66) | 919 (875,963) | -0.10 (-0.35,0.15) | -12 (-40, 17) |
| 5-8w | 0.67 (0.26,1.1) | 44 (22,67) | -0.07 (-0.34,0.2) | -8 (-38,23) |
| 9-12w | 0.50 (0.09,0.91) | 33 (6,60) | -0.14 (-0.44,0.16) | -16 (-51,18) |
| 13-24w | -0.49 (-0.77,-0.21) | -32 (-51,-14) | 0.07 (-0.23,0.38) | 8 (-26,43) |

Beta (B) estimates (95% confidence intervals - CI) represent the difference in the weekly rate (in percentage points) of health care utilization during 1-4 weeks, 5-8 weeks, 9-12 weeks and 13-24 weeks compared with during 12-1 weeks before, for those testing positive for COVID-19 vs the untested comparison group. Estimates compare the post-test period (as indicated) to the pre-test period (12-1 weeks before) for those testing positive for COVID-19, to the same pre- and post-periods for those in the untested comparison group. Categorical variables for utilization in test week, age, sex, calendar month, birth country and comorbidities were included in all models. The % relative diff. transforms the difference-in-difference estimate, which are in percentage points, to the relative change compared with the utilization rate during 12-1 weeks before the PCR test for the untested comparison group (with lower and upper bound of CI also calculated by dividing by the utilization rate during 12-1 weeks before the PCR test for the untested comparison group). PCR: polymerase chain reaction.

We observed no short-term increase in *specialist care* use for any of the age groups following a positive test for SARS-CoV-2, neither at 1-4, 5-8 or 9-12 weeks when compared to untested controls (Table 4).

### Long-term (4-6 months) effects on health care, both comparison groups

When comparing the within-group changes over a longer time span with each other, results were fairly consistent across age groups and comparison groups, generally implying no long-term (13-24 weeks) effects on primary or specialist care use. However, children aged 1-5 years still had a 14% increase in primary care use at 13-24 weeks when compared to same-aged children testing negative (Table 3). No such increase was observed when compared to 1-5-year-old untested children (Table 4). For children aged 6-15 and 16-19 years, we observed no long-term increase in primary- or specialist care after testing positive for SARS-CoV-2, for any of the age groups, independent of whether the comparison group was set to be the untested controls, or the children with a confirmed negative test (Table 3, Table 4).

## Discussion

### Principal findings

In 294 839 children and adolescents tested and 473 721 not tested for SARS-CoV-2 in Norway, we show that children aged 1-19 years have complaints of such a severity that their primary care use, but not specialist care use, is elevated after positive test. The length of increased primary care use depended on age. Children in pre-school age (1-5 years) had a longer elevated use of primary health care services (3-6 months), than 6-19 year-old’s (1-3 months).

To our knowledge, this study is the first study of post-acute effects of COVID-19 in a general child population. By including all children and adolescents tested for SARS-CoV-2 in Norway, we could provide a detailed picture of the health care use for the different age groups. Using routinely collected registry data and applying modern regression techniques, we show that the main impact of COVID-19 on health care use in children and adolescents aged 1-19 years is likely limited to an increase of primary care visits that is of age-dependent length. The findings suggest that COVID-19 does not lead to severe long-term health problems in children and adolescents, at least not problems that require follow-up by (specialist) health care services. These findings are important in the consideration of whether children and adolescents should be vaccinated against SARS-CoV-2 and whether measures to control COVID-19 are still required when all adults have been vaccinated.

### Interpretation & comparison with related studies

An important finding in our study is that the post-covid increase in primary care visits had a longer duration for the younger children than for the older children. More specifically, and when compared to children testing negative, the relative increase in primary care use in the youngest age groups (1-5 years) at 9-12 weeks following positive test, was four times the relative increased in primary care use in the oldest age group (16-19 years) in the same period (Table 3). Also, the 1-5-year-olds’ represented the only age group with a long-term (13-24 weeks) increase in primary care use (Table 3). The trend towards younger children having a longer impact of COVID-19 was less evident in analyses using untested children as a comparison group (Table 4). Thus, altogether, our findings may be summarized by a 3-6 months duration of increased primary care visits for the 1-5-year-olds, a 3 months duration for the 6-15-year-olds, and a 1-3 months duration for the 16-19-year-olds. For all age groups, the increase was mainly due to complaints from the respiratory system (S-Figure 1). Because the youngest patients may experience more long-lasting symptoms also after other respiratory infections^17^, a prolonged increase in primary care use due to respiratory conditions after an infection, as here observed after COVID-19, may be expected.

Our findings shed new light to previous reports showing that COVID-19 may lead to similar post-covid complaints among children as among adults^6, 7^. As an example, a study of 129 children with mean age 11 years reported at least one persisting symptom at 120 days after COVID-19^10^. Typical complaints were fatigue, muscle and joint pain, headache, insomnia, respiratory problems and palpitations^10^. Another study of 171 children with median age 3 years report findings that are more in line with findings in the current study, i.e. that a small proportion (4%) had remaining symptoms 3-8 weeks after the initial infection and that the symptoms were limited to persistent cough and fatigue^11^. In the current study, we show that respiratory complaints and other general/unspecified post-covid complaints are likely to give an increase in health care use, yet it is limited to an increase in visits to the general practitioner. Thus, our findings may imply that the severity grade of post-covid complaints are limited for children (no specialist care needed). These findings of limited impact of mild COVID-19 on long-term morbidities in children, are also consistent with our recent findings of limited long-term comorbidities following mild COVID-19 in adults^1^. However, we here show that the elevation of primary care use may last longer for the youngest children (1-5 years) than what we observed for adults using similar methods^1, 18^.

### Strengths & Limitations

Important strengths of our study are its sample size, the inclusion of children with confirmed positive or negative PCR test for SARS-CoV-2 and the use of two comparison groups. Another strength is the use of routinely collected data from registers that are mandated by law and cover the entire population. This ensures representativeness and that our findings can be used to inform actions to control the virus in Norway and comparable countries. Along this line, the most obvious limitation is that health care utilization cannot always be used as a proxy for a population’s health or medical conditions. Thus, symptoms may persist that are not dealt with in primary or specialist care.

Other important limitations are the methodological challenges arising due to differences in the age-specific testing patterns (Figure 1). As an example, parents may decide to test their 2-year old based on only non-verbal information, whereas teenagers to a larger extent may book a PCR test by own initiative. Although recommendations for testing have included all symptomatic individuals, parents may be more likely to test children having pre-existing conditions and younger children, rather than their healthy and older children, which may give different selection criteria into the different age strata’s groups for those testing positive or negative for SARS-CoV-2. We circumvented this selection issue of the children being at-risk or in need of health care also being more often tested, by three measures. One, we randomly assigned the untested child population – presumably a generally healthy group - a hypothetical test date, which increases the representativity of our study. Two, we excluded all children who had an inpatient or outpatient hospital stay during the test week or the 1-2 weeks following the test week, since such children may be tested as a result of seeking medical treatment. Excluding them ensures that the group testing negative is more similar to the group testing positive in terms of pre- and post-test health care use. And three, we only studied the children who were tested after August 1^st^ 2020, i.e. when testing was widely available for all population groups (not only health care workers and those regarded to be at risk).

Altogether, we believe that the measures taken to combat selection for PCR testing, and the more or less similar findings across two different comparison groups – one possibly healthier (untested) than, and one similarly healthy (no COVID-19), as the children with COVID-19 - as well as across different age groups, strengthen the validity and representativity of our findings. In that regard, our findings are representative for countries that have equal and free access to health care and PCR testing for SARS-CoV-2 for all its inhabitants. Finally, we cannot be certain that the temporal pattern in health care use of children with no COVID-19 or untested are reasonable counterfactuals for the patterns in health care use of the children with COVID-19. However, the similar trends in health care use in the months before the test week (Figure 2) are in line with the common trend assumption of the difference-in-differences method^13^.

### Conclusion

Children aged 1-19 years’ require health care services mainly at the primary care level following a positive test for SARS-CoV-2. No elevated specialist care use was observed following pediatric COVID-19 in the post-acute phase at the population level. The increase in primary care was mainly due to respiratory complaints for all age groups, and children in pre-school age may experience a longer recovery period (∼3-6 months) than children in primary and secondary school age (∼1-3 months).

## Supporting information

ICMJE uniform disclosure form

ICMJE uniform disclosure form

ICMJE uniform disclosure form

ICMJE uniform disclosure form

ICMJE uniform disclosure form

ICMJE uniform disclosure form

ICMJE uniform disclosure form

## Data Availability

Data are not publicly available.

## Acknowledgements

We would like to thank the Norwegian Directorate of Health, in particular Director for Health Registries Olav Isak Sjøflot and his department, for excellent cooperation in establishing the emergency preparedness register. We would also like to thank Gutorm Høgåsen and Anja Elsrud Schou Lindman for their invaluable efforts in the work on the register. The interpretation and reporting of the data are the sole responsibility of the authors, and no endorsement by the register is intended or should be inferred. We would also like to thank everyone at the Norwegian Institute of Public Health who has been part of the outbreak investigation and response team.

## Conflict of interest disclosures

All authors have completed the ICMJE uniform disclosure form and declare: no support from any organization for the submitted work; no financial relationships with any organizations that might have an interest in the submitted work in the previous three years; no other relationships or activities that could appear to have influenced the submitted work.

## Author contribution

Kjetil Telle and Karin Magnusson designed the study. Katrine Damgaard Skyrud, Doris Tove Kristoffersen and Karin Magnusson had access to all of the data in the study and takes full responsibility for the integrity of the data and the accuracy of the data analysis. Katrine Damgaard Skyrud and Doris Tove Kristoffersen performed the statistical analyses and Karin Magnusson drafted the manuscript. Kjetil Telle, Pål Suren, Ketil Størdal and Margrstubethe Greve-Isdahl critically evaluated the stages of the research process. All authors contributed with acquisition of data, conceptual design, analyses and interpretation of results. All authors contributed in drafting the article or critically revising it for important intellectual content. All authors gave final approval for the version to be submitted.

## Funding/support

The study was funded by the Norwegian Institute of Public Health. No external funding was received.

## Role of the funder

The funding sources had no influence on the design or conduct of the study, the collection, management, analysis, or interpretation of the data, the preparation, review, or approval of the manuscript, or the decision to submit the manuscript for publication.

## Supplementary file for the paper

**S-Table 1.**
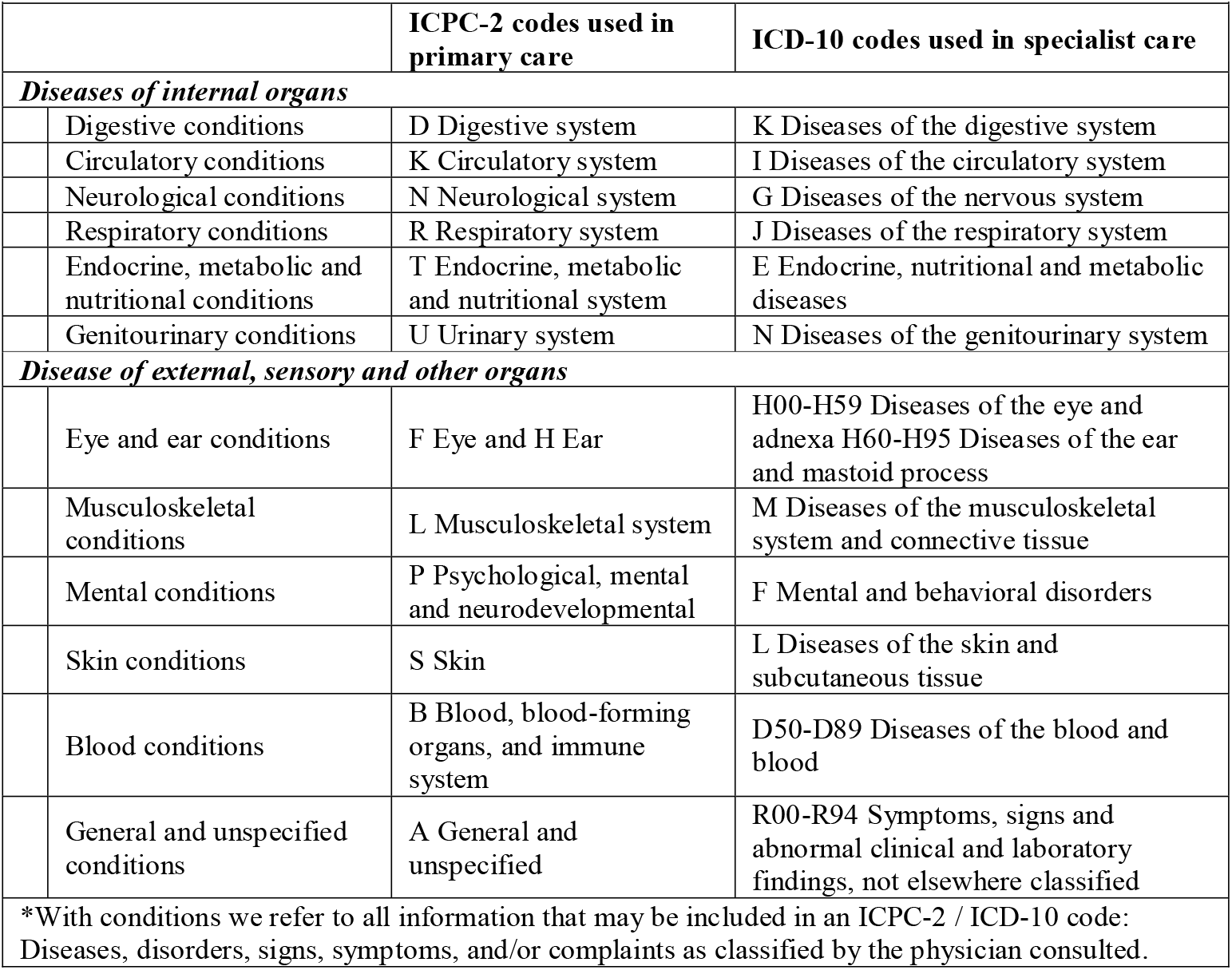
Definitions of the cause-specific diagnosis groups applied.

**S-Figure 1.**
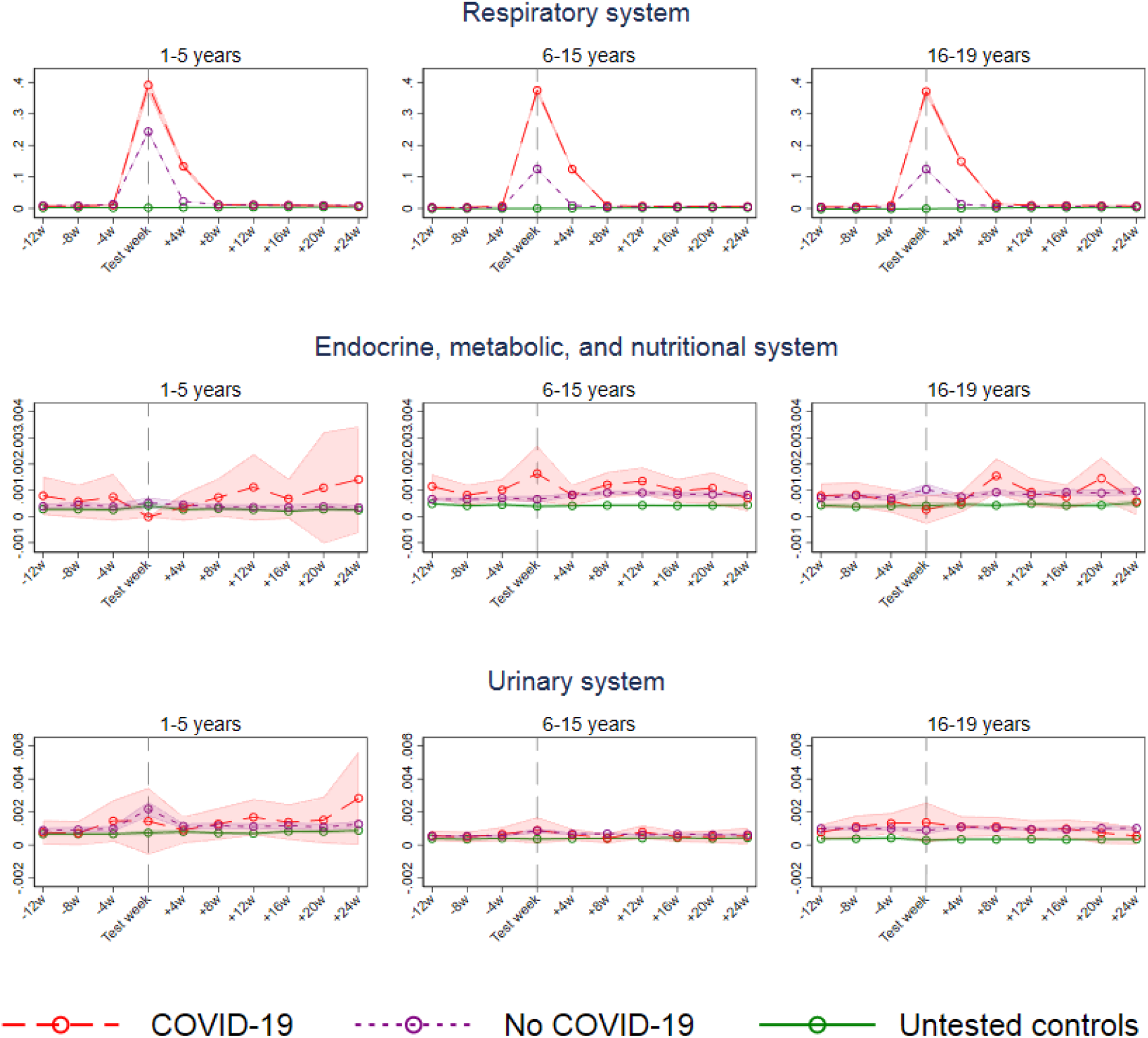
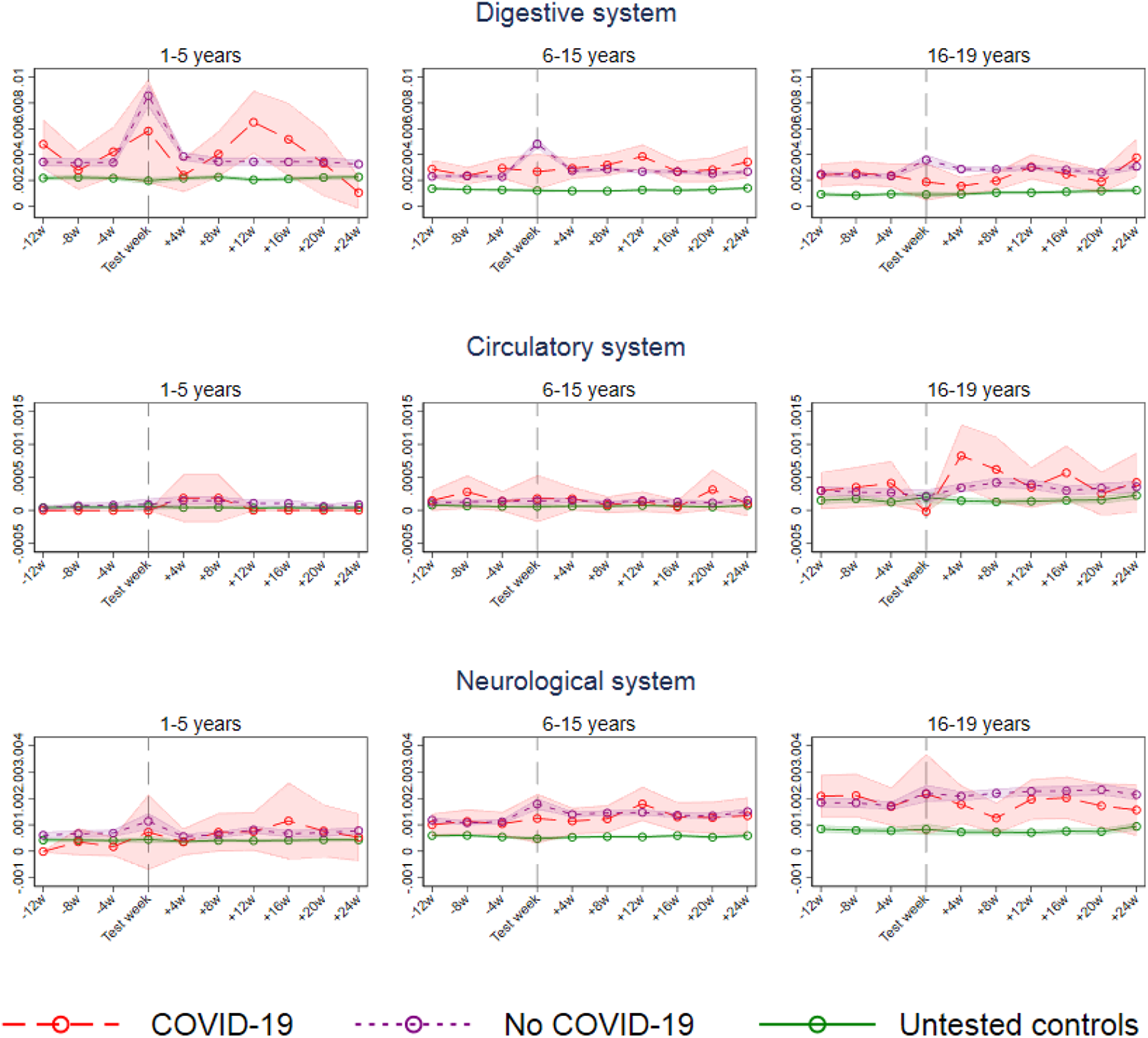
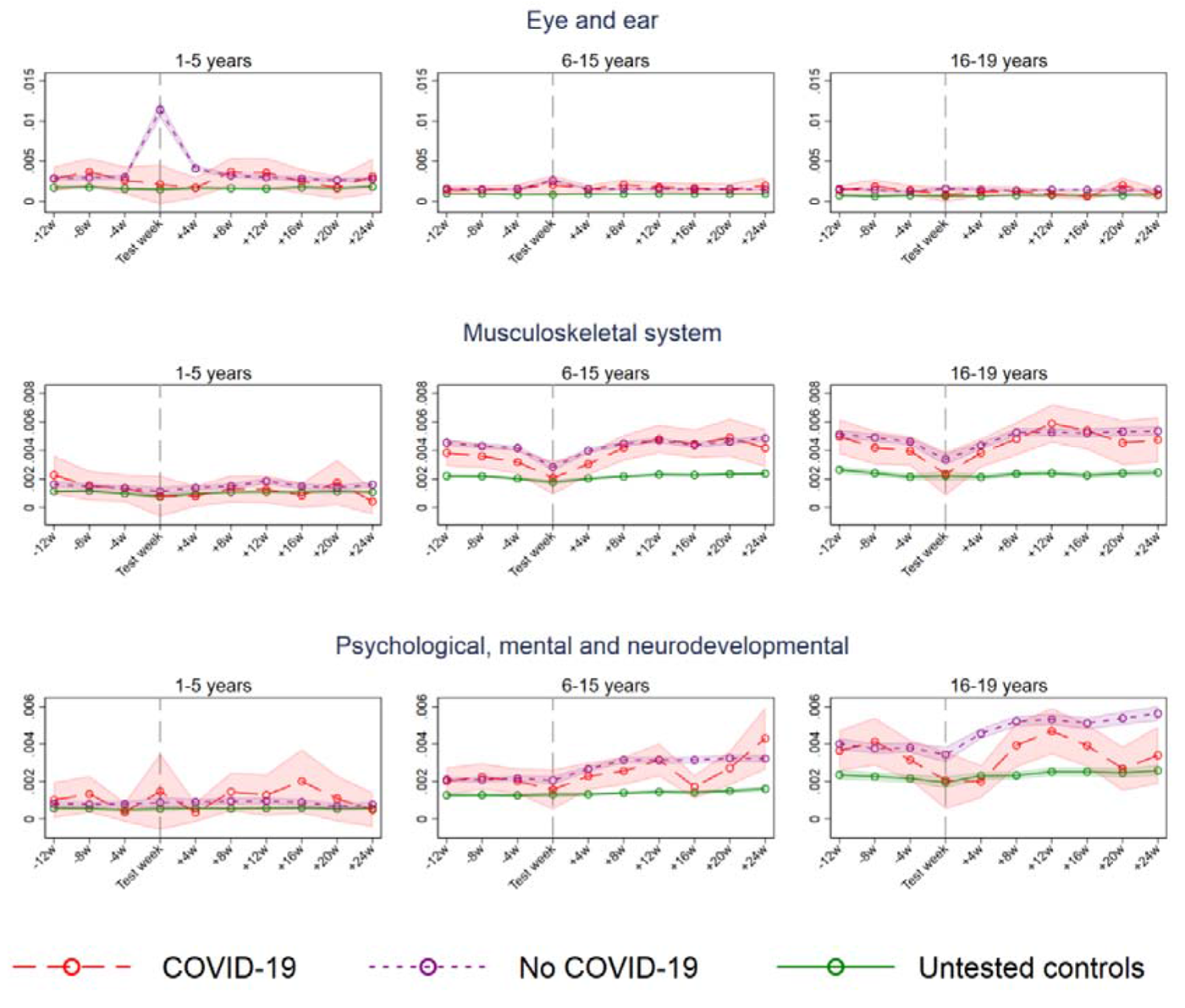
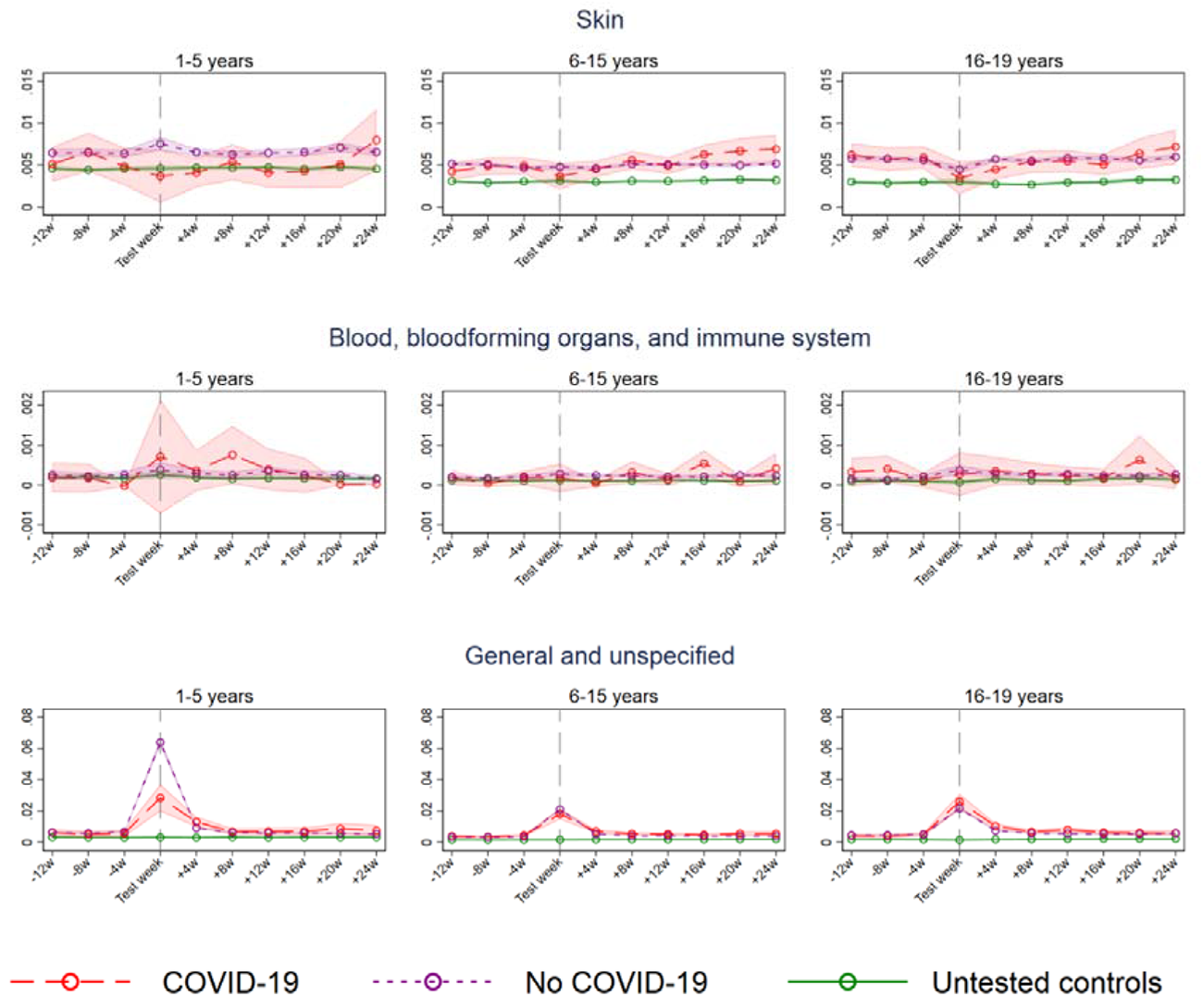
Cause-specific primary health care use for children testing positive and negative and who were untested for SARS-CoV-2 between August 1^st^ 2020 and February 1^st^ 2021.

## Notes

### Competing Interest Statement

The authors have declared no competing interest.

### Author Declarations

Institutional board review was conducted, and The Ethics Committee of South-East Norway confirmed (June 4th 2020, #153204) that external ethical board review was not required.

## References

1. Skyrud KD, Telle KE, Magnusson K. Impacts of COVID-19 on long-term health and health care use. medRxiv 2021.02.16.21251807; doi: https://doi.org/10.1101/2021.02.16.21251807

2. Ayoubkhani D, Khunti K, Nafilyan V, Maddox T, Humberstone B, Diamond I et al. Post-covid syndrome in individuals admitted to hospital with covid-19: retrospective cohort study BMJ 2021; 372:n693 doi:10.1136/bmj.n693

3. Huang, C., Huang L, Wang Y, Li X, Ren L, Gu X, et al., 6-month consequences of COVID-19 in patients discharged from hospital: a cohort study. Lancet 2021. 397(10270): p. 220–232.

4. Lerum TV, Aalokken TM, Bronstad E, Aarli B, Ikdahl E, Lund KMA, et al., Dyspnoea, lung function and CT findings three months after hospital admission for COVID-19. Eur Respir J 2020

5. Himmels JPW, Qureshi SA, Brurberg KG, Gravningen KM. COVID-19: LongTerm Effects of COVID-19 [Langvarige effekter av covid-19. Hurtigoversikt 2021] Oslo: Norwegian Institute of Public Health, 2021.

6. Rubens J H, Akindele N P, Tschudy M M, Sick-Samuels A C. Acute covid-19 and multisystem inflammatory syndrome in children BMJ 2021; 372 :n385 doi:10.1136/bmj.n385

7. Ludvigsson, JF. Systematic review of COVID-19 in children shows milder cases and a better prognosis than adults. Acta Paediatr. 2020; 109: 1088– 1095. https://doi.org/10.1111/apa.15270

8. Ludvigsson, JF. Case report and systematic review suggest that children may experience similar long-term effects to adults after clinical COVID-19. Acta Paediatr. 2021; 110: 914– 921. https://doi.org/10.1111/apa.15673

9. Blomberg B, Mohn K, Brokstad KR, Linchausen D, Hansen BA, Jalloh SL et al. Long COVID affects home-isolated young patients, 23 February 2021, PREPRINT (Version 1) available at Research Square [https://doi.org/10.21203/rs.3.rs-238339/v1]

10. Buonsenso, D., Munblit, D., De Rose, C., Sinatti, D., Ricchiuto, A., Carfi, A. and Valentini, P. (2021), Preliminary evidence on long COVID in children. Acta Paediatr. https://doi.org/10.1111/apa.15870

11. Say D, Crawford N, McNab S, Wurzel D, Steer A, Tosif S. Post-acute COVID-19 outcomes in children with mild and asymptomatic disease. The Lancet Child & Adolescent Health, Volume 5, Issue 6, e22–e23

12. Norwegian Institute of Public Health. The Norwegian Emergency Preparedness Register (BEREDT C19), 2020. ovhttps://www.fhi.no/sv/smittsomme-sykdommer/corona/norsk-beredskapsregister-forcovid-19/

13. Dimick JB, Ryan AM. Methods for Evaluating Changes in Health Care Policy: The Difference-in-Differences Approach. JAMA. 2014;312(22):2401–2402.

14. Angrist J D. Pischke J-S. Most Harmless Econometrics An Empiricist’s Companion. Princeton University Press, 2009.

15. Wing C, Simon K, Bello-Gomez RA. Designing difference in difference studies: best practices for public health policy research. Annual review of public health. 2018;39.

16. Coronavirus immunisation programme in Norway Rational for the recommendations. 2020. Norwegian Insititute of Public Health: Oslo.

17. Bush A. Recurrent Respiratory Infections. Paediatric Clinics of North America, 2009:56;67–100.

18. Skyrud KD, Telle KE, Hernæs KH, Magnusson K. Impacts of COVID-19 on sick leave. medRxiv 2021.04.09.21255215; doi: https://doi.org/10.1101/2021.04.09.21255215

